# Cerebrospinal fluid neurofilament light improves accurate distinction between neurodegenerative and psychiatric disorders at a cognitive neuropsychiatry service

**DOI:** 10.1101/2022.09.08.22279663

**Authors:** Matthew Kang, Dhamidhu Eratne, Hannah Dobson, Charles B Malpas, Michael Keem, Courtney Lewis, Jasleen Grewal, Vivian Tsoukra, Christa Dang, Ramon Mocellin, Tomas Kalincik, Alexander F Santillo, Henrik Zetterberg, Kaj Blennow, Christiane Stehmann, Shiji Varghese, Qiao-Xin Li, Colin L Masters, Steven Collins, Samuel F Berkovic, Andrew Evans, Wendy Kelso, Sarah Farrand, Samantha M Loi, Mark Walterfang, Dennis Velakoulis

## Abstract

**Objective:** People with neuropsychiatric symptoms often experience delay in accurate diagnosis. Although cerebrospinal fluid neurofilament light (CSF NfL) shows promise in distinguishing neurodegenerative disorders (ND) from psychiatric disorders (PSY), its accuracy in a diagnostically challenging cohort longitudinally is unknown.

**Methods:** We collected longitudinal diagnostic information (mean=36 months) from patients assessed at a neuropsychiatry service, categorising diagnoses as ND/mild cognitive impairment/other neurological disorders (ND/MCI/other), and PSY. We pre-specified NfL>582pg/mL as indicative of ND/MCI/other.

**Results:** Diagnostic category changed from initial to final diagnosis for 23% (49/212) of patients. NfL predicted the final diagnostic category for 92% (22/24) of these and predicted final diagnostic category overall (ND/MCI/other vs. PSY) in 88% (187/212), compared to 77% (163/212) with clinical assessment alone.

**Conclusions:** CSF NfL improved diagnostic accuracy, with potential to have led to earlier, accurate diagnosis in a real-world setting using a pre-specified cut-off, adding weight to translation of NfL into clinical practice.

## BACKGROUND

Accurate diagnosis of neuropsychiatric and cognitive presentations can be challenging due to reliance on clinical symptoms, which often overlap between neurodegenerative (ND) and primary psychiatric disorders (PSY).^1,2^ Low mood and memory loss can occur in both Alzheimer’s disease (AD) and late-life depression, creating diagnostic uncertainty.^1^ Moreover, up to 70% of patients with ‘pseudodementia’ subsequently progress to develop ND.^3^ Behavioural variant frontotemporal dementia (bvFTD) is particularly prone to misdiagnosis, as it mimics PSY.^4^

Differentiating between ND and PSY is difficult, resulting in a high rate of misdiagnoses, further compounding the burden of these disorders. A large clinical trial found that 35% of patients with a clinical diagnosis of AD had negative amyloidbeta positron emission tomography scans.^5^ Twenty eight percent of patients at a tertiary memory clinic were diagnosed with PSY before they were eventually rediagnosed with ND, with a mean delay of 33.3 months.^1^ A recent study found that despite multidisciplinary and multimodal assessment, initial ND were subsequently revised to PSY and vice versa for 15% of patients.^4^

To address the challenges associated with clinical diagnoses, international groups have recommended that the diagnosis of dementia should incorporate both clinical and biomarker findings. For example, the recently established Neuropsychiatric International Consortium for Frontotemporal Dementia recommended cerebrospinal fluid (CSF) and blood biomarkers to aid clinicians in reaching an accurate diagnosis.^6^ Similarly, the current conceptual framework for AD incorporates CSF biomarkers in its criteria.^7,8^

Despite the heterogeneity of neurodegenerative and psychiatric disorders, neurofilament light chain (NfL) in CSF has shown promise as a diagnostic discriminator,^9,10^ a finding that our recent studies have replicated.^11,12^ NfL, a neurofilament subunit, make up the structure of the axonal skeleton expressed in large myelinated axons.^13^ Elevated concentrations of NfL can indicate axonal injury and neurodegeneration.^14^ NfL is elevated in other neurological disorders, including inflammatory, traumatic and cerebrovascular disease.^15^ Furthermore, CSF NfL in mild cognitive impairment (MCI), a heterogenous group thought to capture the predementia phase of cognitive decline,^16^ is elevated compared to healthy controls.^14,17^

Previous studies based in clinical settings have assessed whether CSF NfL concentrations correlated with the consensus diagnosis based on cross-sectional multimodal and multidisciplinary assessments.^10,11,18^ However, despite these comprehensive assessments, a significant proportion of patients’ clinical diagnoses are revised longitudinally, including between diagnostic categories (i.e. ND<->PSY).^1,4^ To the best of our knowledge, no studies have examined whether a single CSF NfL test can predict the final diagnostic category in a clinical cohort with longitudinal diagnostic information, with the potential to reduce misdiagnosis for this diagnostically challenging cohort.

This study aimed to determine whether CSF NfL at baseline assessment in a real-world clinical setting could predict the final diagnostic category (ND/MCI/other neurological disorders vs. PSY) using a pre-specified cut-off. We hypothesised that CSF NfL would accurately predict the final diagnostic category. More specifically, a NfL concentration above the pre-specified cut-off would predict a final diagnosis of ND/MCI/other, and NfL below the cut-off would predict a final PSY diagnosis. In instances where there was a diagnostic change (i.e., initial PSY changed to ND/MCI/other, and initial ND/MCI/other changed to PSY), we hypothesised that baseline CSF NfL would have correctly predicted the final diagnostic category. The secondary aim was to identify situations where clinicians should be more cautious in interpreting NfL.

## METHODS

### Study setting

This study was part of The Markers in Neuropsychiatric Disorders Study (The MiND Study; https://themindstudy.org), exploring the use of biomarkers to improve timely diagnosis for people with neuropsychiatric symptoms. This study extends on our previous work, incorporating new patients and updated diagnostic information from longitudinal data.^11,12^

We included 228 patients who had undergone a diagnostic lumbar puncture as part of their baseline assessment at Neuropsychiatry, The Royal Melbourne Hospital, between January 2009 to March 2021 and had sufficient remnant CSF available for NfL analysis. Neuropsychiatry is a tertiary state-wide specialist service providing comprehensive diagnostic assessment of possible dementia, and ongoing management. Included in this study were 201/498 (40.4%) patients from our previous study.^11^

Patients were assessed in inpatient and/or outpatient settings by a multidisciplinary team including neuropsychiatrists, neurologists, neuropsychologists, nurses, occupational therapists, speech pathologists, and social workers, as well as receiving multimodal investigations including brain magnetic resonance imaging (MRI), single-photon emission computerized tomography (SPECT) and fluorodeoxyglucose-positron emission tomography (FDG-PET), and CSF analysis. Patients received consensus diagnosis, with the classification of ND being based on established diagnostic criteria at assessment time.^19–25^ Diagnosis of MCI was based on the Petersen criteria.^26^ PSY diagnoses were defined by the American Psychiatric Association’s Diagnostic and Statistical Manual of Mental Disorders (4^th^ Edition; Text Revision [DSM-IV-TR]^27^ and 5^th^ Edition [DSM-5]^28^).

### Data collection

Three authors (Kang, Dobson, Keem) who were not involved with the clinical assessments at baseline and were blinded to the NfL concentration performed a file review using a codebook to ensure internal reliability, including discharge summaries and outpatient assessment reports. The data included demographic information, clinical variables, and the initial diagnosis from the baseline assessment at the patient’s presentation to the service. For patients with follow-up internally within Neuropsychiatry, we used subsequent multidisciplinary assessment letters to collect their final diagnosis. For patients who were discharged from Neuropsychiatry, we contacted the primary doctor still involved in the patient’s care (e.g., private neurologist/psychiatrist, mental health clinic, family physician, other specialty clinic) and asked them to provide the final diagnosis (blinded to NfL). A minimum of 12-months of follow-up information was available for all participants.

We grouped initial and final diagnoses into categories:

***Neurodegenerative disorders (ND)*** included all dementias and other disorders associated with neurodegeneration (e.g., Parkinson’s disease).

***MCI/other -*** MCI, considered a pre-dementia entity,^16^ and other neurological disorders (i.e., epilepsy and acquired brain injury) were categorised as another group.

***Primary psychiatric disorders (PSY)*** included psychoses, mood disorders, anxiety disorders, post-traumatic stress disorder, alcohol/substance use disorder, functional neurological/cognitive disorders and psychiatric-mixed (where there were multiple psychiatric diagnoses).

***Diagnosis unclear -*** a small proportion of patients were diagnostically unclear despite comprehensive assessment, with further interval assessment being scheduled.

### Cerebrospinal fluid measurements

CSF analysis has been previously described.^11^ Briefly, CSF was stored at –80°C, and NfL measured using a commercial enzyme-linked immunosorbent assay (ELISA; NF-light; UmanDiagnostics, Sweden, distributed by abacus dX), according to the manufacturer’s protocols, at the National Dementia Diagnostic Laboratory, Melbourne. CSF was diluted 1+1 and reconstituted standards were added to the plate in duplicate, incubated, and washed. Samples displaying concentrations above the highest standard point were further diluted and re-assayed. Two internal controls of pooled CSF were also included in each NfL plate. Mean intra- and inter-assay Coefficient of variation for NfL was 6.2% and 11.3% respectively.^29^

### NfL concentration cut-off

Although various cut-offs for CSF NfL have been reported, generalisability is limited, as NfL concentrations vary depending on the lab performing analysis.^15,30^ We used a pre-specified cut-off (582pg/mL), based on our previous study which demonstrated a 95% positive predictive value (PPV) and 78% negative predictive value (NPV).^11^

NfL was rated as accurately predicting a final ND/MCI/other or PSY when: a) the patient’s final diagnosis was ND/MCI/other and their baseline concentration was greater than 582pg/mL, or b) the patient’s final diagnosis was PSY and their baseline concentration was less than 582pg/mL.

### Ethics

This study was approved by Human Research Ethics Committees at Melbourne Health (2016.038, 2017.090, 2018.371, 2020.142) and University of Melbourne (1341074, 1648441.3).

### Statistical analysis

Statistical analyses were performed using Jamovi and R Studio.^31–35^ Continuous variables were reported as means with standard deviations. Categorical variables were reported as frequencies (%). As a result of the smaller sample in the diagnostically unstable group, Mann-Whitney U tests were performed to test differences in numerical variables between diagnostically stable and unstable groups. Pearson’s chi-square tests of independence were performed to compare dichotomous variables and determine differences in psychiatric and family history. A two-sided p-value<0.05 was considered statistically significant. General linear models (GLMs) were estimated to compare the differences in NfL between the diagnostic categories using age at CSF sampling as a covariate.

Bias-corrected and accelerated confidence intervals (CIs) were computed for all GLM and ROC parameters via non-parametric bootstrapping with two thousand replicates. Statistical significance was defined as any CI not capturing the null-hypothesis value (at the 95% level). These robust statistical methods were selected because they mitigate the effects of distributional violations, including the presence of outliers.

Given the heterogeneity and small sample size of patients of MCI/other diagnoses and our previous finding^11^ that patients with MCI had an intermediary NfL concentration (n=5, 739pg/mL, 95%CI[478, 1001]) compared to ND (1528pg/mL, 95%CI[1168, 1888]) and PSY (435pg/mL, 95%CI[394,476]), we also performed analyses comparing ND vs. PSY.

As a post-hoc analysis, we calculated different cut-offs for participants in older (>=60 years old) and younger groups (<60 years old) using receiver operator characteristic (ROC) curves. Area under the ROC curve (AUC), sensitivity, and specificity of NfL in distinguishing ND from PSY were computed. Optimal cut-off was determined using Youden’s J statistic.

## RESULTS

### Study Cohort (Table 1)

Of 228 eligible patients, 13 were lost to follow-up, and three died within one year following their baseline assessment (AD, Lewy body dementia/DLB and progressive supranuclear palsy). The remaining 212 patients had a mean age of 55.4 years (SD 11.6), 41% (87/212) were female, and the mean follow-up period was 34.2 months (SD 24.1, range 12-148), which was similar among the final diagnostic groups.Participants with a follow-up diagnosis of PSY were younger than the ND and MCI/other group (mean difference of 8.2 and 8.8 years respectively).

**Table 1.**
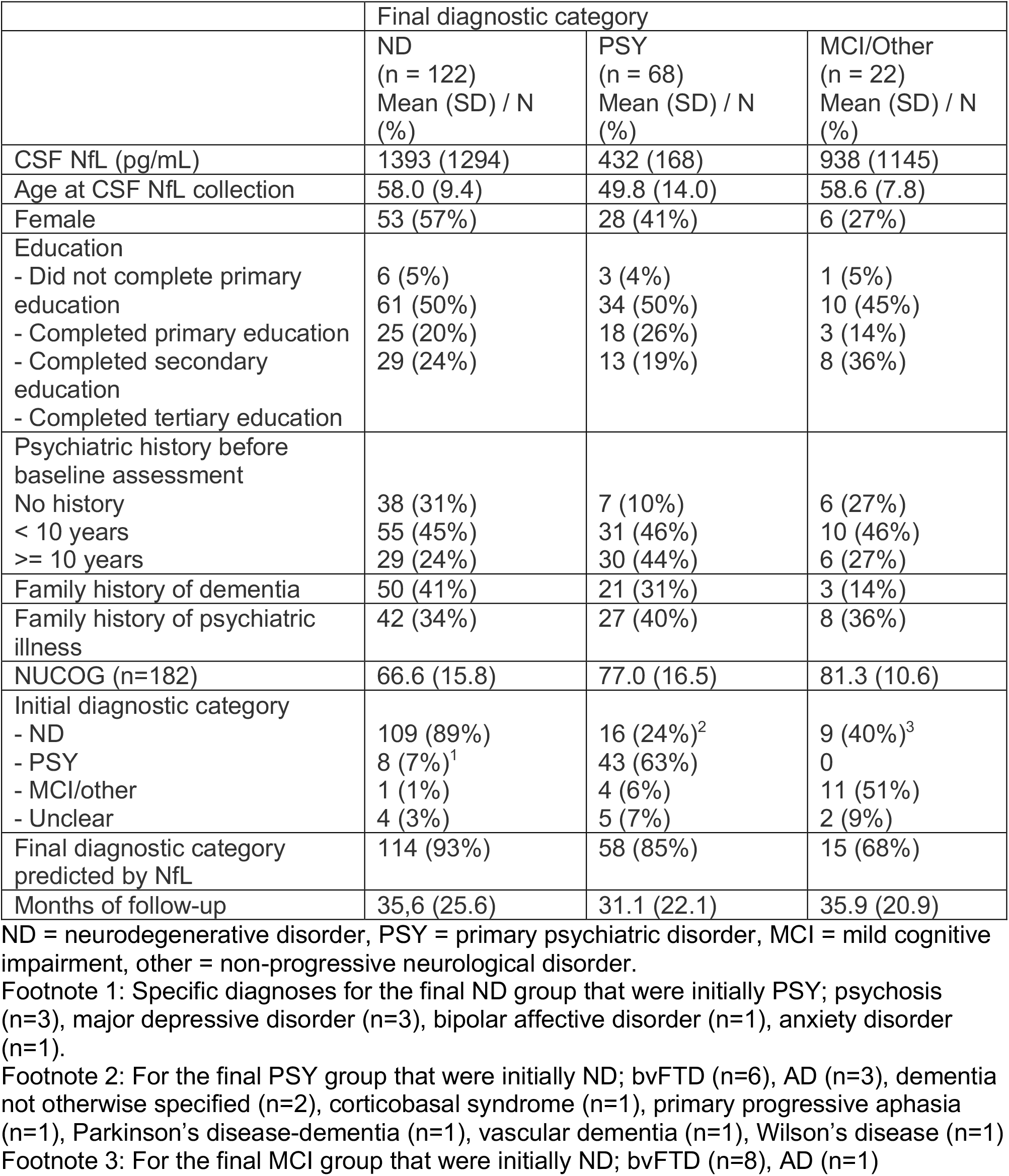
Participant demographics, final diagnostic category and NfL accuracy.

Adjusting for age, baseline CSF NfL concentrations were higher in patients with a final diagnosis of ND compared to PSY (GLM, mean difference=929pg/mL, 95%CI:[598, 1259]). Concentrations in the MCI/other group were higher than PSY (mean difference=449pg/mL, CI:[156, 743]) but lower than ND (mean difference=428pg/mL, CI:[-157.8, 1014]). These are presented in Figure 1.

**Figure 1.**
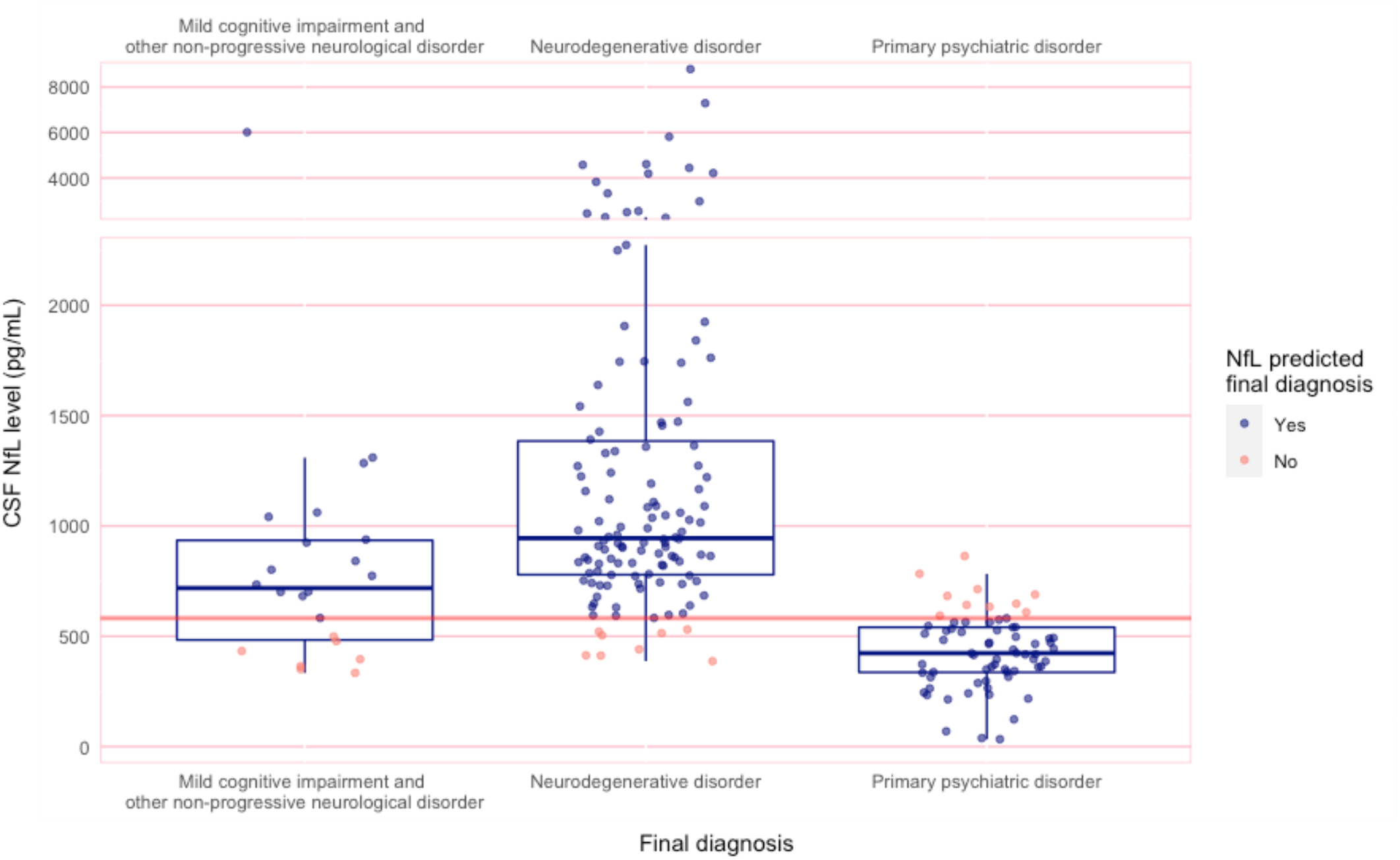
Boxplot of NfL levels in diagnostic groups.

### ND group

Fifty-eight percent (122/212) of patients had a final diagnosis of ND, including AD (n=51), frontotemporal lobar degeneration (FTLD n=24, bvFTD n=16), dementia not otherwise specified (n=13), corticobasal syndrome (CBS, n=7), vascular dementia (n=6), DLB (n=4), mixed dementia (n=3), Parkinson’s disease (n=3, and others including Niemann-Pick Type C, Huntington’s disease, multiple sclerosis, and CNS vasculitis. Four patients initially deemed as being diagnostically unclear at baseline assessment eventually received a ND diagnosis (AD, CBS, DLB, and mixed dementia).

#### MCI and other neurological illnesses

Ten percent (22/212) of patients had a final diagnosis of MCI/other. There were 18 with MCI, two with epilepsy, one with acquired brain injury, and one whose neurocognitive symptoms completely resolved without any comorbid psychiatric illnesses.

#### PSY group

Thirty-two percent (68/212) participants had a final PSY diagnosis, including psychosis (n=23), bipolar disorder (n=6), major depression (n=9), functional neurological disorder (n=6), obsessive-compulsive disorder (n=1), post-traumatic stress disorder (n=1), and mixed psychiatric disorders (n=18). Of five patients deemed diagnostically unclear at baseline assessment, all received a final diagnosis of PSY, which were depression (n=2), FTD phenocopy (n=1), and psychiatric-mixed (n=2).

### Diagnostic change from initial diagnosis at baseline assessment to final diagnosis

A visual representation of the participants’ diagnostic stability is shown in Figure 2. There was a change in diagnostic category from initial to final diagnosis for 23% (49/212) patients. bvFTD was the most frequently changed diagnosis (14/49, 29%). Of note, 48 patients from our previous publication^11^ had their diagnosis category changed and updated with the incorporation of the new longitudinal data of this study.

**Figure 2.**
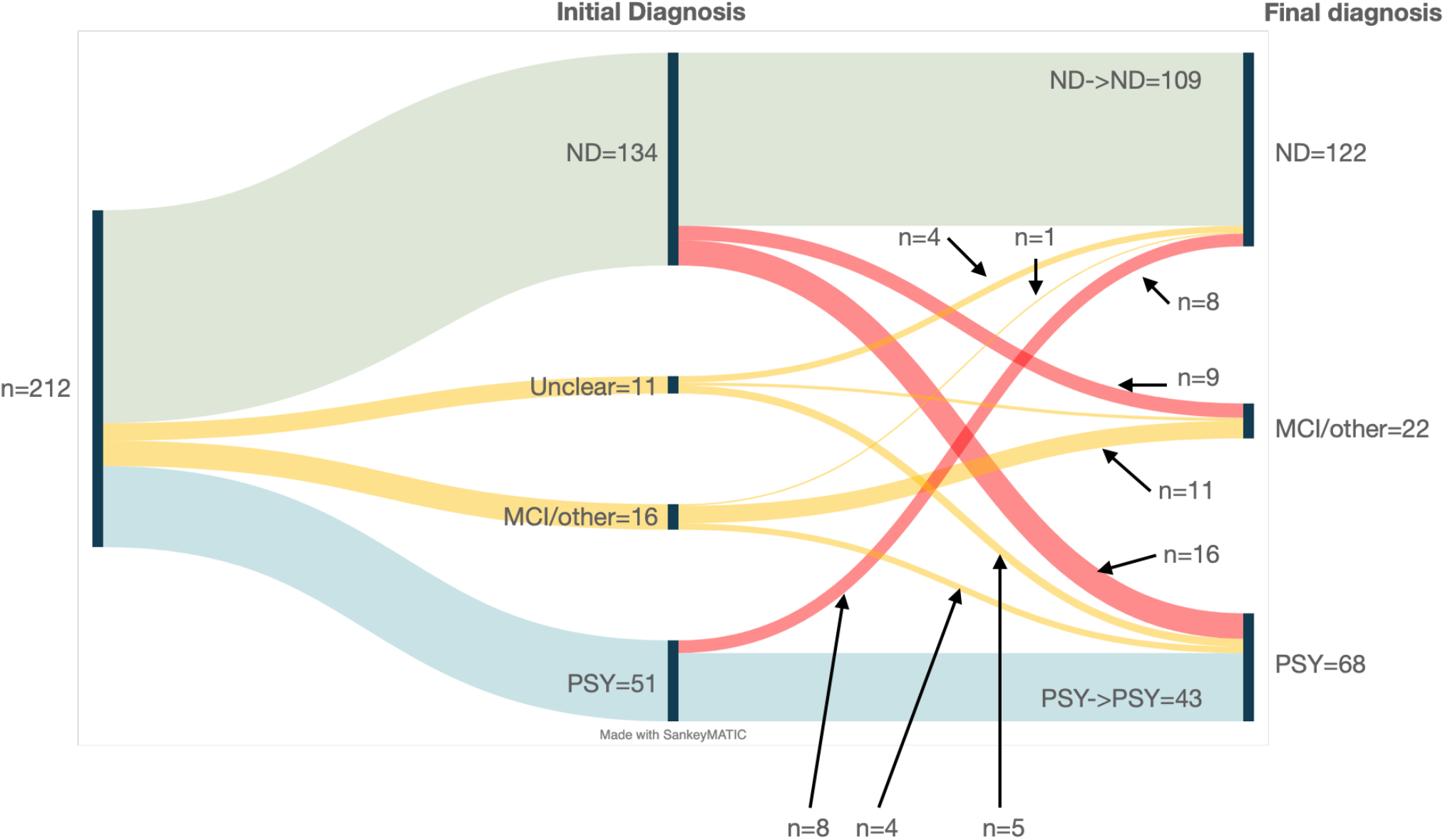
Sankey diagram of diagnostic journey. MCI/other: Mild cognitive impairment/other non-progressive neurological disorder ND: Neurodegenerative disorder PSY: Primary psychiatric disorder Unclear: Diagnostically unclear at initial assessment

Twenty-four patients (11%, 24/212) patients had changed between ND and PSY as shown in Table 2. Eight patients (8%, 8/122) who were given a final diagnosis of ND were initially diagnosed with PSY at their baseline diagnosis before it was revised (PSY->ND). Sixteen patients (24%, 16/68) who were given a final diagnosis of PSY were initially diagnosed with ND (PSY->ND). Half (9/18) of the MCI patients initially had baseline diagnoses of ND.

**Table 2.** Comparison of those with diagnostic category stability vs change.

|  | Change in diagnosis |  | No change in diagnosis |  |
| --- | --- | --- | --- | --- |
|  | PSY->ND | ND->PSY | ND-> ND | PSY->PSY |
| N | 8 | 16 | 109 | 43 |
| Baseline CSF NfL | 1040 (SD 503) | 401 (SD 114) | 1435 (SD 1351) | 426 (SD 185) |
| Predicted by CSF NfL | 6 (75%) | 16 (100%) | 104 (95.4%) | 36 (83.7%) |
| Age at CSF NfL collection | 63.1 (SD 8.41) | 52.9 (SD 10.9) | 57.3 (SD 9.38) | 48.6 (SD 15.1) |
| Female | 4 (50%) | 8 (50%) | 43 (42.2%) | 17 |
| Education |  |  |  |  |
| - Did not complete primary education | 1 | 0 | 4 | 2 |
| - Completed primary education | 6 | 5 | 54 | 25 |
| - Completed secondary education | 1 | 6 | 22 | 11 |
| - Completed tertiary education | 0 | 5 | 28 | 5 |
| Psychiatric history before baseline assessment |  |  |  |  |
| No history | 0 | 1 | 37 | 5 |
| < 10 years | 2 | 7 | 50 | 20 |
| >= 10 years | 6 | 8 | 22 | 18 |
| Family history of dementia | 4 | 13 | 65 | 27 |
| Family history of psychiatric illness | 66 | 8 | 70 | 28 |
| NUCOG (n=149) | 52.6 (16.1) | 72.1 (17.8) | 67.2 (15.8) | 78.8 (16.5) |
| Initial diagnosis | Anxiety 1<br>BPAD 1<br>Depression 2<br>PSY-other 1<br>Psychosis 3 | AD 3<br>bvFTD 6<br>CBD 1<br>Dementia NOS 2<br>Other neurodeg 1<br>PD 1<br>PPA 1<br>VD 1 | AD 50<br>bvFTD 20<br>CBD 3<br>DLB 4<br>Dementia NOS 13<br>MS 1<br>Mixed dementia 1<br>Other neurodeg 9<br>PD 3<br>SD 3<br>VD 2 | BPAD 5<br>Depression 5<br>FND 5<br>OCD 1<br>PTSD 1<br>PSY-other 10<br>Psychosis 16 |
| Months of follow-up | 48.9 (40.7) | 41.1 (27.3) | 34.4 (24.3) | 28.3 (21.2) |

AD – Alzheimer's disease, BPAD – bipolar affective disorder, bvFTD – behaviour variant frontotemporal dementia, CBD – corticobasal degeneration, DLB – dementia with Lewy bodies, Dementia NOS – Dementia not otherwise specified, FND – functional neurological disorder, MS – multiple sclerosis, OCD – obsessive-compulsive disorder, PD – Parkinson's disease, PPA – primary progressive aphasia, PSY-other – other primary psychiatric disorder, PTSD – post-traumatic stress disorder, SD – semantic dementia, VD – vascular dementia,

A comparison of clinical variables of patients that changed between ND and PSY is in Table 2. Patients who changed diagnostic categories (ND->PSY or PSY->ND) were more likely to have a long psychiatric history (>10 years, n=14/22, x_2_=10.6, p=0.005). None of the other clinical variables were statistically different.

A table detailing description of the patients with change in their diagnostic category can be found in the supplementary file.

### Accuracy of CSF NfL in predicting the final diagnosis category

#### Final diagnosis

CSF NfL at baseline assessment predicted the final diagnostic category (ND/MCI/other vs. PSY) for 88% patients (187/212). The accuracy was higher for predicting ND (93%) and PSY (85%), compared to MCI/other (68%), as shown in Table 1.

NfL improved the diagnostic accuracy for all diagnostic categories (ND, MCI/other and PSY) compared to initial diagnosis from the baseline assessment, as shown in Figure 3.

**Figure 3.**
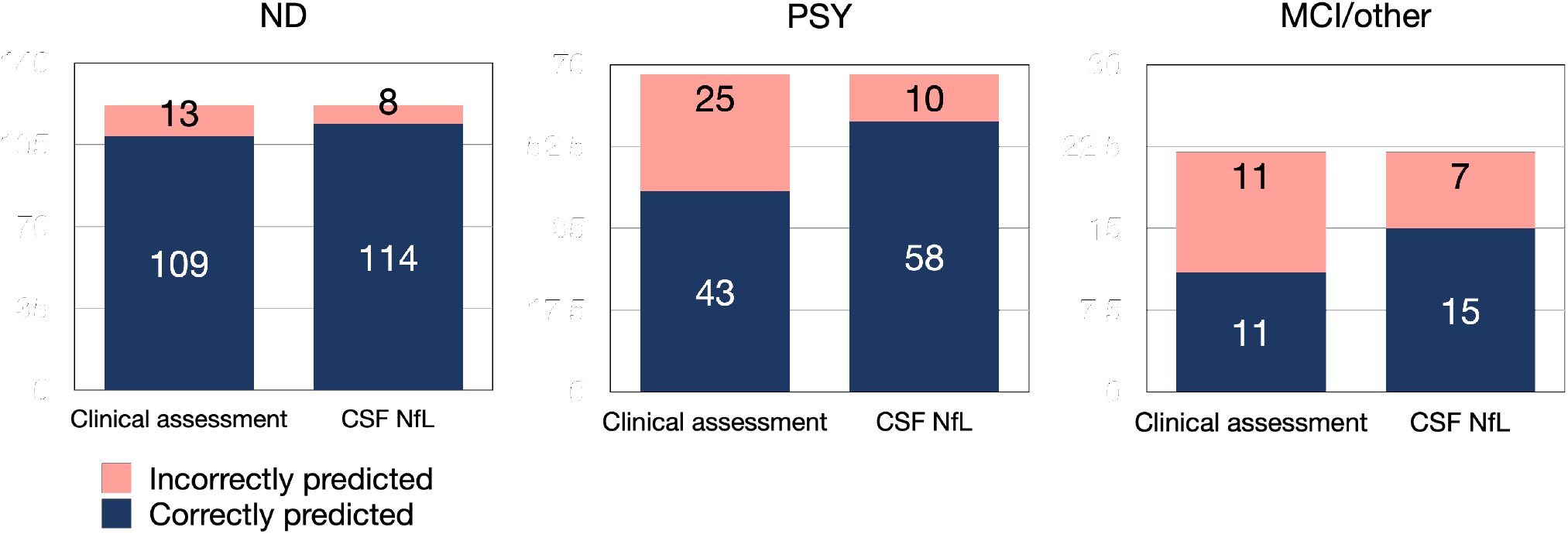
Accuracy of baseline CSF NfL in predicting final diagnosis versus initial clinical assessment. CSF NfL at baseline accurately predicted 114/122 (93%) of final ND diagnoses compared to 109/122 (89%) baseline clinical assessments. For PSY the rates were 58/68 (85%) vs 43/25 (63%), and for MCI/Other the rates were 15/22 (68%) vs 11/22 (50%) for CSF NfL and baseline clinical assessments respectively MCI/other = mild cognitive impairment and other non-progressive neurological disorders, ND = neurodegenerative disorders, PSY = primary psychiatric disorders

For the 24 patients whose diagnostic category changed between ND and PSY, NfL accurately predicted the final diagnostic category for 92% (22/24). The two patients where the baseline NfL did not predict the final diagnosis were both PSY->ND and, notably, their follow-up duration between their initial and final diagnosis was 90 and 100 months respectively, compared to the other 22 patients (mean=38.5 months, SD 27.6).

Of the nine who were initially diagnostically unclear at their baseline assessment and then had a final diagnosis of PSY or ND diagnosis, 78% (7/9) were predicted by their baseline NfL. Four of the five (80%) patients initially diagnosed with MCI before they were given a final diagnosis of PSY or ND were predicted by their NfL.

When comparing NfL to only the initial diagnosis from the baseline assessment (i.e., before longitudinal follow-up and the final diagnosis after changes/refinement), its apparent accuracy was only 79% (159/201).

### Age-specific cut-offs

We determined optimal CSF NfL cut-offs for distinguishing ND/MCI/other from PSY, in people younger than 60 years and 60 and above (Figure 4). The optimal cut-off for <60 years was the same as our pre-determined cut-off (582pg/mL). The optimal cut-off for >=60 years was 716pg/mL.

**Figure 4.**
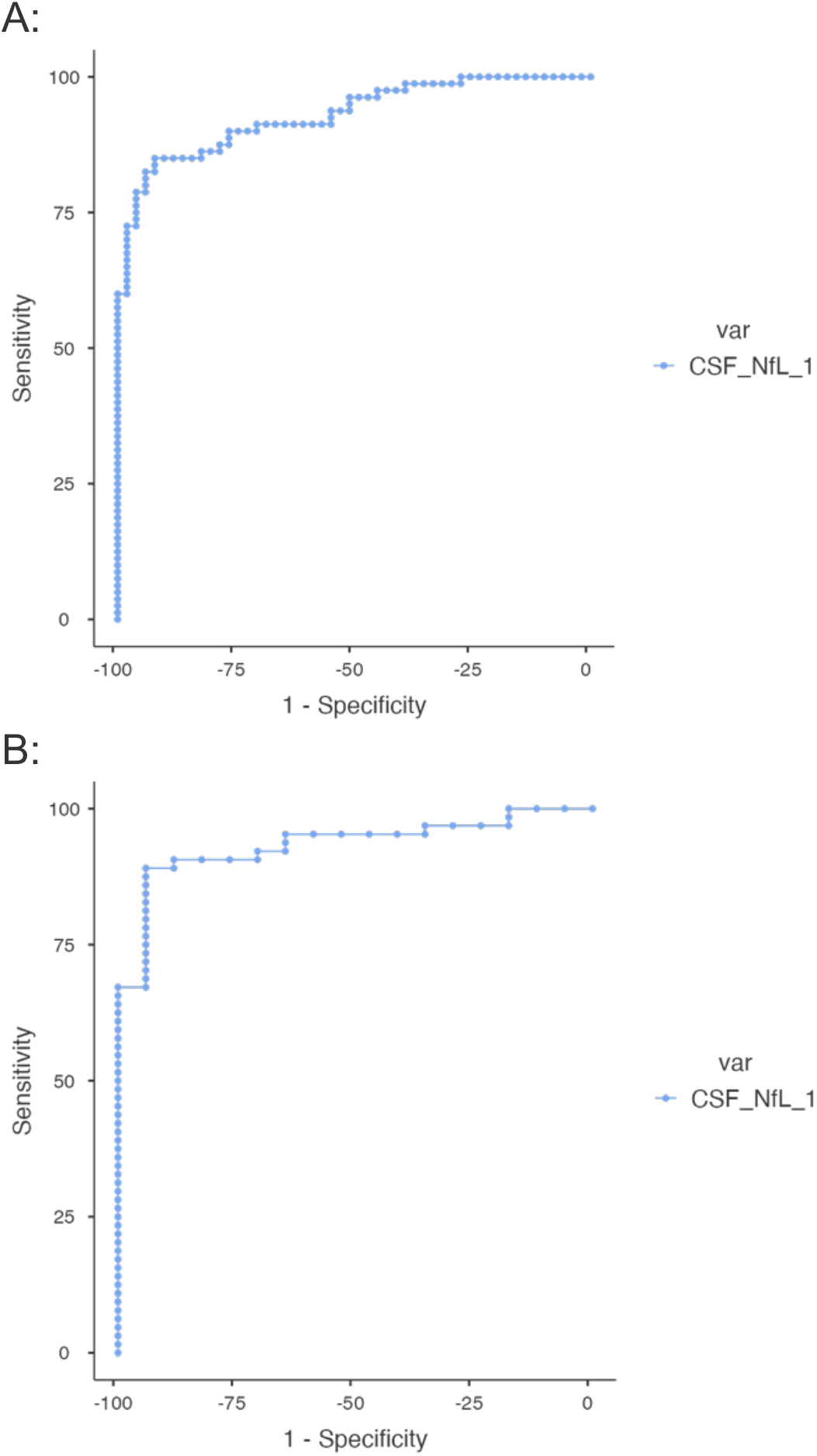
ROC curves for patients <60 years of age (A) and 60+ years of age (B)

Using this 716pg/mL cut-off for the 60+ cohort improved accuracy of baseline CSF NfL in predicting final ND/MCI/other vs PSY diagnoses from 90% (64/71) to 93% (66/71). The number of people 60 years and older was small (n=81, ND=54, PSY=17, MCI/other=17); therefore, results for the older group must be interpreted with caution.

## DISCUSSION

We found that a single CSF NfL test could have accurately predicted a final diagnosis of a neurodegenerative disorder or MCI/other neurological disorder, versus a primary psychiatric, for 88% of cases in a real-world, inclusive, clinical cohort assessed for cognitive and neuropsychiatric presentations. Furthermore, we found that CSF NfL, in retrospect, could have aided clinicians in avoiding a delayed diagnosis for 11% of patients compared to relying on baseline clinical diagnostic assessment alone. Our finding of a change in diagnostic category longitudinally in 23% of patients based on gold standard assessments in a tertiary cognitive neuropsychiatry service is consistent with the literature on the challenges faced in clinical practice.^1,4^ The potential for NfL to improve diagnostic accuracy supports the translation of NfL from research labs into routine patient care. This is in line with recent recommendations from international groups to use CSF biomarkers such as NfL to assist in challenging clinical distinctions such as differentiating AD and bvFTD from psychiatric and non-neurodegenerative causes.^6–8^

Strengths of this study include the generalisable cohort, inclusion of all patients regardless of comorbidities, longitudinal follow-up and diagnoses based on serial multimodal and multidisciplinary assessments, and blinded comprehensive review of files and follow-up. This study builds on previous work with new longitudinal diagnostic information^11,12^ demonstrating that CSF NfL could assist clinicians in ruling in or out psychiatric disorders being the cause of a patient’s neuropsychiatric presentation with diagnostic stability over time. To our knowledge, no other studies have examined the diagnostic utility of NfL to predict and reduce diagnostic delay in such an inclusive, real-world clinical cohort, with a focus on younger people where misdiagnosis and diagnostic delay are more common.^36^ In particular, we saw that bvFTD was the most frequently changed diagnosis, and our study adds further support to the utility of biomarkers in this challenging diagnosis.^6^

We found that the accuracy of CSF NfL in predicting the diagnosis improved (88% final diagnosis with longitudinal follow-up vs 79% initial diagnosis from baseline assessment) when longitudinal diagnostic information was incorporated (mean follow-up period=34.2 months, SD 24.1), This is consistent with clinical practice, where diagnoses based on clinical assessments are often revised with serial assessments and time. It highlights the value of longitudinal follow-up information and updated diagnoses. Therefore, it is possible that studies that relied only on current diagnostic criteria without longitudinal information may have underestimated the accuracy of NfL compared to this study.

Our findings where baseline CSF NfL was not consistent with the final diagnosis (i.e., baseline NfL<=582pg/mL but final ND, or >582pg/mL but final PSY) highlight scenarios where clinicians should interpret NfL with caution. This was in the cases where patients had Parkinson’s disease, comorbid cerebrovascular burden, and MCI. The small number of patients with Parkinson’s disease had a low baseline NfL concentration despite Parkinson’s disease being a progressive neurodegenerative disorder. This is in line with previous findings that suggest NfL, found primarily in myelin-rich axons, may not be affected in Parkinson’s disease where there is no widespread axonal degeneration.^15^ Patients with a final diagnosis of PSY but with evidence of chronic small vessel disease on their neuroimaging may still have a mildly elevated NfL concentration. However, their NfL concentrations were within 20% of the cut-off and not as markedly increased as concentrations seen in ND group. This may be NfL detecting psychiatric disorder associated with white matter hyperintensities, including ‘vascular depression’.^37,38^ Future studies characterising the relationship between NfL in patients with primary psychiatric disorders and chronic small vessel disease in their brain will help inform clinicians on how to interpret NfL in this context. NfL concentrations were higher in those with MCI compared to patients diagnosed with PSY, consistent with previous literature.^14,39^ MCI is a complex group to characterise due to the heterogeneous nature of their cognitive difficulties, with variable use in clinical and research setting.^16^ As CSF NfL has been linked to the progression rate of neurodegenerative disorders,^30^ it may in turn indicate the prognosis of those with MCI and whether the condition will progress to definitive neurodegenerative disorder, such as AD. Furthermore, there may be a role for serial CSF NfL in these higher-risk individuals, as the trend of NfL may help indicate whether there is a neurodegenerative or static process that may be too slow or subtle for other biomarkers such as neuroimaging to be able to detect.

Strengths of this study include the generalisable cohort, inclusion of all patients regardless of comorbidities, longitudinal follow-up and diagnoses based on serial multimodal and multidisciplinary assessments, and blinded comprehensive review of files and follow-up information. While a strength of our study was using a pre-determined cut-off, this cut-off may need to be adjusted for other services due to variabilities between labs.^15,30^ Therefore, ongoing international work to develop reference ranges for research and clinical use, such as the Global Biomarker Standardization Consortium initiatives,^40^ is critical. Furthermore, our findings support age-specific cut-offs to increase diagnostic accuracy, and diagnostic performance may be higher in younger people with cognitive and neuropsychiatric presentations. These age-specific cut-offs require validation in an external cohort to ensure that the classification accuracy is robust. In addition, more recent studies examining blood NfL in multiple sclerosis used age-adjusted z-scores to mitigate the non-linear relationship between NfL age,^41^ which was not possible for this study due to the lack of a normative dataset for CSF NfL, but could be a consideration for future studies.

Limitations of this study included those inherent to a retrospective design, mitigated by longitudinal follow-up data collection and categorisation, and blinded, rigorous, and comprehensive reviews. Despite obtaining several years of follow-up with serial assessments for most patients, more insights may be yielded with longer follow-up and/or post-mortem and pathological confirmation. For example, there may be some participants diagnosed with psychiatric disorders, who may instead have an extremely slowly progressing neurodegenerative illness. In these cases, we may have underestimated the number of neurodegenerative disorders. Studies exploring the diagnostic and wider utility of serial NfL are underway.

This study showed that CSF NfL could be a useful diagnostic tool to help differentiate neurodegenerative disorders from psychiatric disorders in clinical practice. This could function not too dissimilar to C-reactive protein in infective/inflammatory conditions. An elevated NfL could help dismiss psychiatric differentials and potential misdiagnosis. Conversely, a low NfL could (especially if persistently low) reduce the risk of a neurodegenerative disorder misdiagnosis and increase confidence in a psychiatric diagnosis. A borderline NfL could prompt careful review of other clinical features and follow-up, and potentially help with prognosticating in complex situations like MCI. While results from this study show potential diagnostic utility of CSF NfL, lumbar punctures are impractical for widespread use in primary care settings. Blood NfL, which correlates well with CSF NfL,^42^ would be more accessible, potentially as a first-tier test as part of a routine ‘dementia panel’ for general practitioners. Therefore, a study investigating the utility of blood NfL in participants recruited much earlier (i.e., at referral) and in primary care and community settings is underway. This single test could reduce misdiagnosis and diagnostic delay many patients and their families face, facilitating earlier precision care and treatment.

## Data Availability

All data produced in the present study are available upon reasonable request to the authors

## ACKNOWLEDGEMENTS AND FUNDING SOURCES

The authors would like to thank all the staff, past and present, at Neuropsychiatry, The Royal Melbourne Hospital, and all the clinicians who provided follow up clinical information.

Finally, the authors would like to thank all the patients and their families for their participation.

We are grateful for funding that supported this work: the Trisno Family Research Grant in Old Age Psychiatry, three NorthWestern Mental Health Research Seed Grants, MACH MRFF, and NHMRC (1185180).

Alexander F Santilli has been supported specifically for this project by The Fromma Foundation, The Ellen and Henrik Sjöbring Foundation, and the Fredrik and Ingrid Thuring Foundation.

Henrik Zetterberg is a Wallenberg Scholar supported by grants from the Swedish Research Council (#2018-02532), the European Research Council (#681712 and #101053962), Swedish State Support for Clinical Research (#ALFGBG-71320), the Alzheimer Drug Discovery Foundation (ADDF), USA (#201809-2016862), the AD Strategic Fund and the Alzheimer’s Association (#ADSF-21-831376-C, #ADSF-21-831381-C and #ADSF-21-831377-C), the Bluefield Project, the Olav Thon Foundation, the Erling-Persson Family Foundation, Stiftelsen för Gamla Tjänarinnor, Hjärnfonden, Sweden (#FO2022-0270), the European Union’s Horizon 2020 research and innovation programme under the Marie Skłodowska-Curie grant agreement No 860197 (MIRIADE), the European Union Joint Programme –

Neurodegenerative Disease Research (JPND2021-00694), and the UK Dementia Research Institute at UCL (UKDRI-1003).

Kaj Blennow is supported by the Swedish Research Council (#2017-00915), the Alzheimer Drug Discovery Foundation (ADDF), USA (#RDAPB-201809-2016615), the Swedish Alzheimer Foundation (#AF-930351, #AF-939721 and #AF-968270), Hjärnfonden, Sweden (#FO2017-0243 and #ALZ2022-0006), the Swedish state under the agreement between the Swedish government and the County Councils, the ALF-agreement (#ALFGBG-715986 and #ALFGBG-965240), the European Union Joint Program for Neurodegenerative Disorders (JPND2019-466-236), the National Institute of Health (NIH), USA, (grant #1R01AG068398-01), and the Alzheimer’s Association 2021 Zenith Award (ZEN-21-848495).

AHE reports honoraria for presentations from Merck, Allergan, Ipsen, Teva, UCB, Abbott, AbbVie, Pfizer, STADA. Participation in scientific advisory board meetings with Allergan, AbbVie, Ipsen, Pfizer and STADA. He holds shares in GKC and CSL.

The corresponding author had full access to all the data in the study and had final responsibility for the decision to submit for publication.

## DECLARATION OF INTERESTS AND FINANCIAL DISCLOSURES

No declarations for:

- Matthew Kang
- Dhamidhu Eratne
- Christa Dang
- Jasleen Grewal
- Michael Keem
- Alexander Santillo
- Vivian Tsoukra
- Hannah Dobson
- Wendy Kelso
- Ramon Mocellin
- Christiane Stehmann
- Shiji Varghese
- Qiao-Xin Li

Steven Collins has acted as a paid consultant to Biogen Australia.

Henrik Zetterberg: HZ has served at scientific advisory boards and/or as a consultant for Abbvie, Acumen, Alector, ALZPath, Annexon, Apellis, Artery Therapeutics, AZTherapies, CogRx, Denali, Eisai, Nervgen, Novo Nordisk, Passage Bio, Pinteon Therapeutics, Red Abbey Labs, reMYND, Roche, Samumed, Siemens Healthineers, Triplet Therapeutics, and Wave, has given lectures in symposia sponsored by Cellectricon, Fujirebio, Alzecure, Biogen, and Roche, and is a co-founder of Brain Biomarker Solutions in Gothenburg AB (BBS), which is a part of the GU Ventures Incubator Program (outside submitted work).

Charles Malpas has received conference travel support from Merck, Novartis, and Biogen. He has received research support from the National Health and Medical Research Council, Multiple Sclerosis Research Australia, The University of Melbourne, The Royal Melbourne Hospital Neuroscience Foundation, and Dementia Australia.

Kaj Blennow has served as a consultant, at advisory boards, or at data monitoring committees for Abcam, Axon, BioArctic, Biogen, JOMDD/Shimadzu. Julius Clinical, Lilly, MagQu, Novartis, Ono Pharma, Pharmatrophix, Prothena, Roche Diagnostics, and Siemens Healthineers, and is a co-founder of Brain Biomarker Solutions in Gothenburg AB (BBS), which is a part of the GU Ventures Incubator Program, outside the work presented in this paper.

Mark Walterfang has served as a consultant, at advisory boards, or on data monitoring committees for Actelion, Vtesse, Sucampo, Mallinckrodt, and Biomarin Pharmaceuticals; has received investigator grants from Pfizer, Lilly, Bristol Meyers Squibb, Actelion, Vtesse, and Mallinckrodt pharmaceuticals; the National Health and Medical Research Council, the Royal Melbourne Hospital, the Ara Parseghian Medical Research Foundation, the Bethlehem Griffiths Foundation, and the CHDI Initiative. He has received travel support from Lilly, Pfizer, Actelion, Orphan, Biomarin, Vtesse and Sucampo Pharmaceuticals.

Tomas Kalincik served on scientific advisory boards for MS International Federation and World Health Organisation, BMS, Roche, Janssen, Sanofi Genzyme, Novartis, Merck and Biogen, steering committee for Brain Atrophy Initiative by Sanofi Genzyme, received conference travel support and/or speaker honoraria from WebMD Global, Eisai, Novartis, Biogen, Roche, Sanofi-Genzyme, Teva, BioCSL and Merck and received research or educational event support from Biogen, Novartis, Genzyme, Roche, Celgene and Merck.

Sarah Farrand has received honoraria from Abbvie, Abbott and a fellowship from Medtronic.

## Notes

### Author Declarations

Human Research Ethics Committees at Melbourne Health gave ethical approval for this work (2016.038, 2017.090, 2018.371, 2020.142) Ethics Committee at University of Melbourne gave ethical approval for this work (1341074, 1648441.3).

